# The Generalizability of Clinical Prediction Models for Patients with Acute Coronary Syndromes: Results from Independent External Validations

**DOI:** 10.1101/2021.01.21.21250234

**Authors:** Benjamin S. Wessler, Jason Nelson, Jinny G. Park, Hannah McGinnes, Jenica Upshaw, Ben Van Calster, David van Klaveren, Ewout Steyerberg, David M. Kent

## Abstract

**Purpose:** It is increasingly recognized that clinical prediction models (CPMs) often do not perform as expected when they are tested on new databases. Independent external validations of CPMs are recommended but often not performed. Here we conduct independent external validations of acute coronary syndrome (ACS) CPMs.

**Methods:** A systematic review identified CPMs predicting outcomes for patients with ACS. Independent external validations were performed by evaluating model performance using individual patient data from 5 large clinical trials. CPM performance with and without various recalibration techniques was evaluated with a focus on CPM discrimination (c-statistic, % relative change in c-statistic) as well as calibration (Harrell’s E_avg_, E_90_, Net Benefit).

**Results:** Of 269 ACS CPMs screened, 23 (8.5%) were compatible with at least one of the trials and 28 clinically appropriate external validations were performed. The median c statistic of the CPMs in the derivation cohorts was 0.76 (IQR, 0.74 to 0.78). The median c-statistic in these external validations was 0.70 (IQR, 0.66 to 0.71) reflecting a 24% decrement in discrimination. However, this decrement in discrimination was due mostly to narrower case-mix in the validation cohorts compared to derivation cohorts, as reflected in the median model based c-statistic [0.71 (IQR 0.66 to 0.75). The median calibration slope in external validations was 0.84 (IQR, 0.72 to 0.98) and the median E_avg_ (standardized by the outcome rate) was 0.4 (IQR, 0.3 to 0.8). Net benefit indicates that most CPMs had a high risk of causing net harm when not recalibrated, particularly for decision thresholds not near the overall outcome rate.

**Conclusion:** Independent external validations of published ACS CPMs demonstrate that models tested in our sample had relatively well-preserved discrimination but poor calibration when externally validated. Applying ‘off-the-shelf’ CPMs often risks net harm unless models are recalibrated to the populations on which they are used.

## Introduction

Ischemic heart disease remains the leading cause of death worldwide and acute coronary syndrome (ACS) accounts for the majority of this morbidity and mortality.^1^ ACS encompasses a spectrum of diseases with different treatment approaches^2–4^ and for individual patients short- and long-term risks of adverse outcomes vary substantially based on clinical characteristics.^5,6^ It is now well recognized that risk-sensitive treatment decisions are relevant across the continuum of care for patients with ACS: whether to pursue an early invasive strategy^7^ (and by what approach^8^), intra-procedural medication use^9^, discharge from the hospital^10^ and duration of long-term medical treatments.^11^ Since risk prediction has become central to contemporary care, it is critical to objectively assess the performance of ACS clinical predictive models (CPMs).

Despite the proliferation of CPMs for cardiovascular disease, many of these CPMs have not been externally validated.^12^ For the CPMs that have been externally validated, there are often major decrements in performance.^13,14^ These observations are a threat to the trustworthiness of predictions as poorly-calibrated models may motivate harmful decisions.^15^ To our knowledge, the performance of available CPMs for ACS has not been extensively evaluated. Additionally, the value of alternative updating procedures that allow CPMs to fit better to new populations has not been examined.

Here, using data from the Tufts PACE CPM Registry, we perform independent external validations of ACS CPMs using individual patient-level data from publically available clinical trials and evaluate model performance with and without updating.

## Methods

### General Approach

We matched available CPMs predicting outcomes after ACS to publically available patient-level data from trials of ACS interventions. For each CPM/ database match, model performance was assessed before and after recalibration.

### Systematic Review of CPMs

This study examined data from the Tufts Predictive Analytics and Comparative Effectiveness (PACE) CPM Registry (http://pace.tuftsmedicalcenter.org/cpm). The Registry has been previously reported^12^ and represents a systematic review of CPMs for patients at risk for and with known cardiovascular disease (CVD). Briefly, PubMed was searched for English-language articles containing CPMs for CVD published from January 1990 through 2015 (search terms available in Supplement). For inclusion in the Registry, articles must (1) develop a CPM as a primary aim, (2) contain at least 2 outcome predictors, and (3) present enough information to estimate the probability for an individual patient. The analysis presented here focuses on CPMs predicting outcomes for patients with ACS.

### Validation Databases

The National Heart, Lung, and Blood Institute (NHLBI) makes de-identified patient-level data available via the Biologic Specimen and Data Repository Information Coordinating Center (BioLINCC) website. Data from trials representing patients who had ACS were requested and screened for inclusion.

### CPM/Database Matching Procedure

The matching process used a tiered approach (Figure 1). Potential matches were first screened by non-clinical staff for inclusion based on crude matching (overlap) of the index condition (STEMI, NSTEMI, unstable angina) and outcomes (including secondary outcomes from the validation clinical trials databases). This process was repeated by clinical experts. CPMs/ validation database pairs that were deemed potentially relevant were then reviewed at a granular level to identify whether the variables included in the CPM were available in the validation database and if the predicted outcome was assessed.

**Figure 1.**
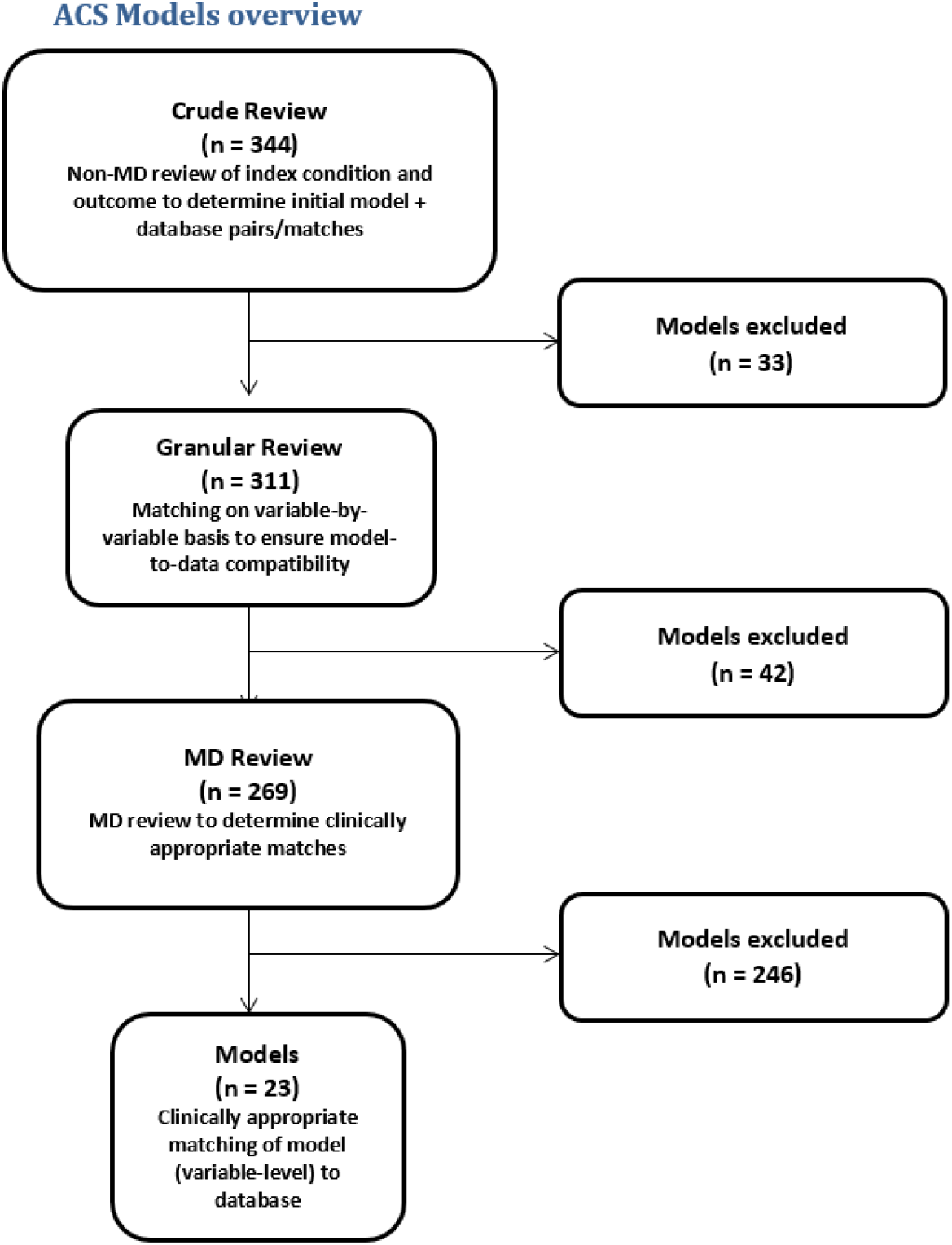

### Relatedness

Because clinical populations even with the same index condition may differ substantially from one another, and because a substantial decrement in performance might be anticipated when models are tested on databases substantially different from those on which they are derived, we assessed the similarity between the CPM derivation population and validation population using a relatedness rubric that classified matches into three categories—“related”, “distantly related” or “no match”. The relatedness rubric is show in the Supplement. For each potential match the derivation and validation populations were assessed across the following domains: Inclusion criteria, exclusion criteria, recruitment setting, revascularization approach, enrollment years, follow up time and outcome. Matches were classified as “related” when there were no major differences across these domains as determined by a clinical expert. Matches were “distantly related” if there was incomplete overlap across these domains (e.g. similar clinical populations treated with different revascularization strategies). Potential matches were excluded as “no match” if the index condition or outcome was sufficiently dissimilar so as to make the application of the CPM clinically inappropriate. The “related” and “distantly related” matches are shown in the Supplement.

### Measuring CPM performance

Using patient-level data from the validation databases, the linear predictor was calculated using the originally published beta coefficients and intercept for each patient. Observed outcomes in the patient-level data were defined using the CPM outcome definition and prediction time horizon. For all model validations observed outcome events that occurred after the prediction time horizon were censored. For time-to-event models, Kaplan Meier estimator was used for right censored follow up times. For binary outcome models, unobserved outcomes (i.e. due to loss-to-follow up prior to the prediction time horizon) were considered missing and excluded from analyses.

*CPM discrimination* was evaluated using the c-statistic. The primary assessment presented here is the change in c-statistic. The percent change in discrimination was calculated as [(Validation c-statistic - 0.5) – (Derivation c-statistic - 0.5) / (Derivation c-statistic - 0.5) * 100].^14^ Discriminatory performance was also assessed by comparing the measured discrimination in the validation database to the model-based concordance measure (MB-c)^16^. The MB-c yields the c-statistic that would be obtained in the validation database under the assumption that the CPM is perfectly valid in the validation database. Consequently, a difference between the derivation c-statistic and the MB-c is fully due to a difference in case-mix between the derivation and validation database. It thus provides a useful benchmark to understand how much the observed changes in discrimination can be attributed to changes in case mix heterogeneity vs model validity.

*Model Calibration* was assessed by converting the linear predictor (*lp*) to the estimated event probability using the equation [predicted value = (1/ (1+e^-*lp*^))]. *Calibration-in-the-large*, a measure of global fit, as well as *calibration slope* and *Harrell’s E* (E_avg_) standardized to the outcome rate were assessed. E_avg_ computes the average absolute calibration error (difference between the observed outcome rate and the estimated probabilities, where the observed rate is estimated using a non-parametric locally weighted scatterplot smoothing). E_90_ representing the 90^th^ percentile of the absolute calibration error was also calculated. Clinical utility was summarized using *net benefit*^17^ for 3 arbitrarily chosen decision thresholds: the outcome prevalence rate, at prevalence/2, and 2*prevalence. Net benefit integrates model performance (calibration and discrimination) with information about the relative utility weights of false-positive and false-negative predictions (as implicitly determined by the decision threshold) to provide a more comprehensive assessment of the potential clinical consequences of using CPMs to inform treatment decisions. We used standardized net benefit^18^ to make results comparable across validations. For this analysis net benefit of 0 was considered neutral (no benefit or harm).

### CPM Recalibration

CPM recalibration was done with three separate techniques. The first recalibration method addresses calibration-in-the-large. In this approach the difference between the mean observed outcome rates in the derivation and validation cohorts was used to update the intercept. The second recalibration technique corrected the intercept (as in the first method) and also applied a uniform correction factor to the regression coefficients to better fit the validation cohort (i.e. correction of the intercept and slope). The third approach to CPM recalibration re-estimates the regression coefficients to better fit the validation database.

## Results

### Validation Databases

Patient level data from 5 large randomized trials were identified and used as validation databases for the ACS CPMs. These trials were AMIS^19^, TIMI-II^7^, TIMI-III^20^, MAGIC^21^, ENRICHD^22^, and detailed trial data compared to CPM populations are shown in Supplement. ≥ 98% of observations in each of these trials were used for the validations presented here. The number of censored events that occurred beyond the CPM prediction time horizon are shown in the Supplement. The Aspirin Myocardial Infarction Study (AIMS) was a multicenter, randomized, double-blind, placebo-controlled trial of aspirin therapy (1 gram) given to patients who had a myocardial infarction. The first patient was randomized in 1975. A total of 4,524 patients were enrolled and followed for 36 months. The primary outcome of coronary heart disease mortality occurred at similar rates in the placebo and aspirin treatment arms. The Thrombolysis in Myocardial Infarction (TIMI) phase II trial was multicenter randomized trial of early invasive vs. conservative management of patients with STEMI treated with recombinant tissue plasminogen activator (TPA). 3534 patients had been enrolled by 1988 and at 6 weeks the primary outcome of reinfarction or death occurred in 10.9% of the invasive strategy group and 9.7% of the conservative group (P not significant). The TIMI III trial was a 2×2 factorial multi-center trial comparing TPA vs placebo as initial strategy and also early invasive vs early conservative strategy for patients with unstable angina and non-Q-wave myocardial infarction. 1473 patients were enrolled between 1989 and 1992. At 6 weeks the primary outcome of death, myocardial infarction, or failure of initial therapy occurred in 54.2% of the TPA-treated patients and 55.5% of the placebo treated patients (P not significant). For the comparison of early invasive vs. conservative management strategies the outcome of death, myocardial infarction or unsatisfactory symptom-limited exercise stress test at 6 weeks occurred in 18.1% of patients in the conservative strategy arm and 16.2% of the early invasive arm (P not significant). The Magnesium in Coronaries (MAGIC) Trial was a randomized multi-center double-blind trial of IV magnesium sulphate or placebo administered to STEMI patients. At 30-days there was no difference in the primary outcome of all-cause mortality between groups (OR 1.0, 95% CI 0.9-1.2). The Enhanced Recovery in Coronary Heart Disease (ENRICHD) multicenter trial was conducted from 1996 to 2001 and randomized 2481 patients with myocardial infarction and depression to cognitive behavior therapy-based psychosocial intervention or usual medical care. At 6 months the primary outcome of death or recurrent MI occurred in 24.1% of patients in the usual care arm and 24.2% of the intervention arm (HR 1.01, 95% CI 0.86-1.18).

### CPM Validation Matching

269 clinically appropriate ACS CPMs were identified (Figure 1). 246 models (91.4%) of these models could not be matched to the available validation databases because of missing variables or non-overlapping outcomes. 23 (8.6%) ACS CPMs were matched at the granular level (match for both variables and outcome on an appropriate clinical population) to the 5 publically available clinical trial databases (Supplement). 18 CPMs were matched to a single database. 5 CPMs were matched to 2 databases. In total there were 28 CPM/ validation database matches.

### CPM Discrimination in Independent External Validations

The median derivation c-statistic was 0.76 (IQR, 0.74 to 0.78) and the median validation c-statistic was 0.70 (IQR, 0.66 to 0.71) representing a 24% loss in discriminatory ability (Table 1). For validations done on “related populations” there was a median 19% loss in discriminatory ability (IQR, −27% to −9%). For validations done on “distantly related” populations there was a median 29% loss (IQR, −44% to −15%) in performance (Figure 1). However, the median MB-c was 0.71 (0.66-0.75) for these CPMs, very similar to the measured c-statistic on the validation databases, indicating that the narrower case mix of the validation population accounted for most of the decrement Models had moderately better preserved discrimination when tested on “related” compared to “distantly-related” databases (Figures 2 and 3).

**Table 1.**

| (n = 28) | Mean (SD) | Median (IQR) | Range |
| --- | --- | --- | --- |
| ***Discrimination (0.76 in development)*** |  |  |  |
| Validation Model Based-c | 0.71 (0.05) | 0.71 (0.66, 0.75) | 0.61, 0.8 |
| Validation c-statistic | 0.69 (0.05) | 0.7 (0.66, 0.71) | 0.58, 0.77 |
| % change (vs development) | -25 (23) | -24 (-37, -15) | -73, 45 |
| % change (vs MB-c) | -5 (30) | -13 (-28, 7) | -50, 72 |
| ***Calibration (8.3% observed outcome rate)*** |  |  |  |
| Slope | 0.85 (0.29) | 0.82 (0.72, 0.95) | 0.24, 2 |
| *standardized E* | 0.6 (0.5) | 0.4 (0.3, 0.8) | 0.1, 2.5 |
| *standardized E90* | 1.4 (1.6) | 1 (0.6, 1.5) | 0.2, 8.4 |
| ***Net Benefit (standardized to prevalence)*** |  |  |  |
| sNB prevalence / 2 | -0.03 (0.14) | 0 (-0.02, 0.04) | -0.5, 0.08 |
| sNB prevalence | 0.23 (0.10) | 0.25 (0.19, 0.29) | 0.01, 0.48 |
| sNB prevalence * 2 | 0.02 (0.15) | 0.02 (0, 0.11) | -0.42, 0.23 |

**Figure 2.**
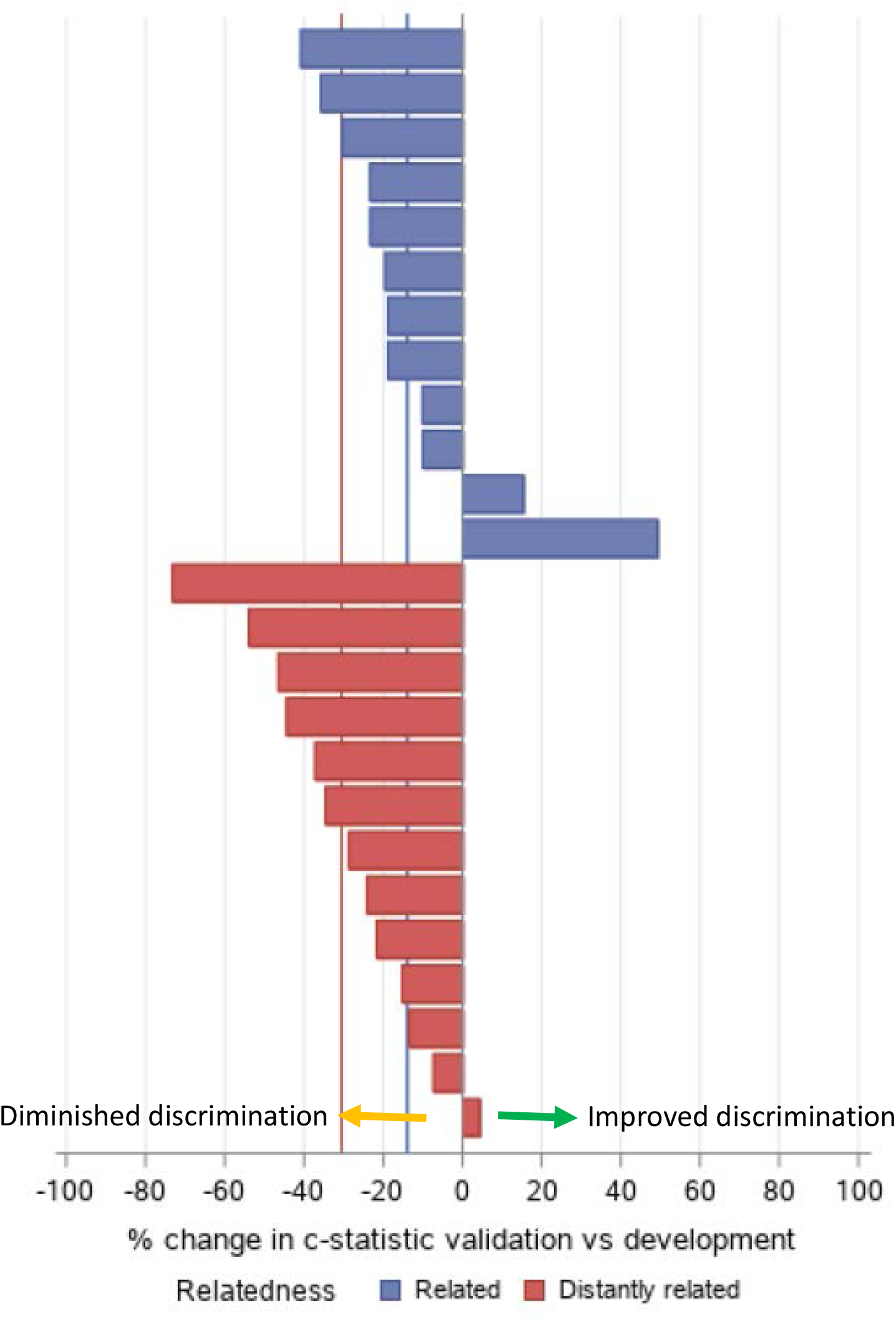

**Figure 3.**
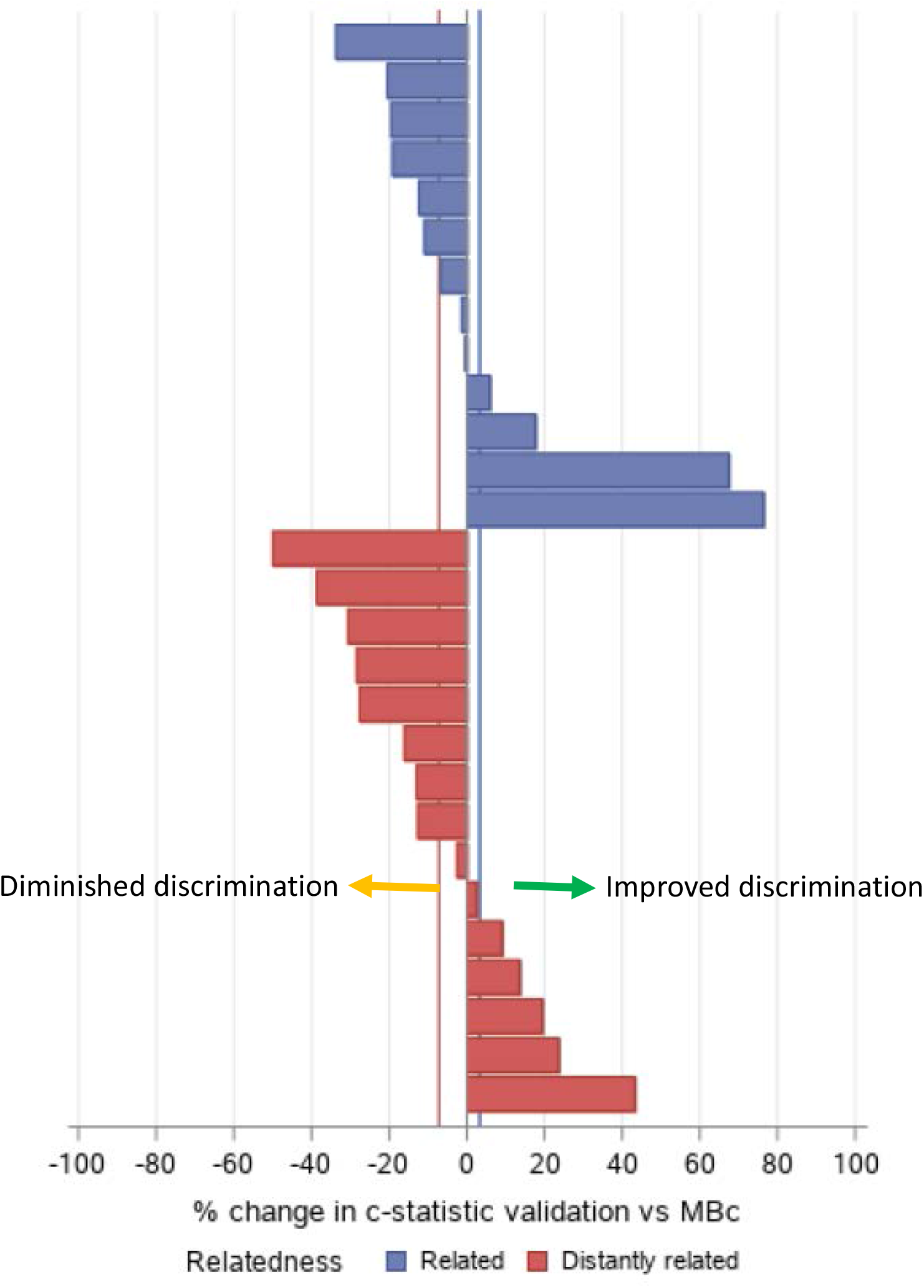

**Figure 4.**
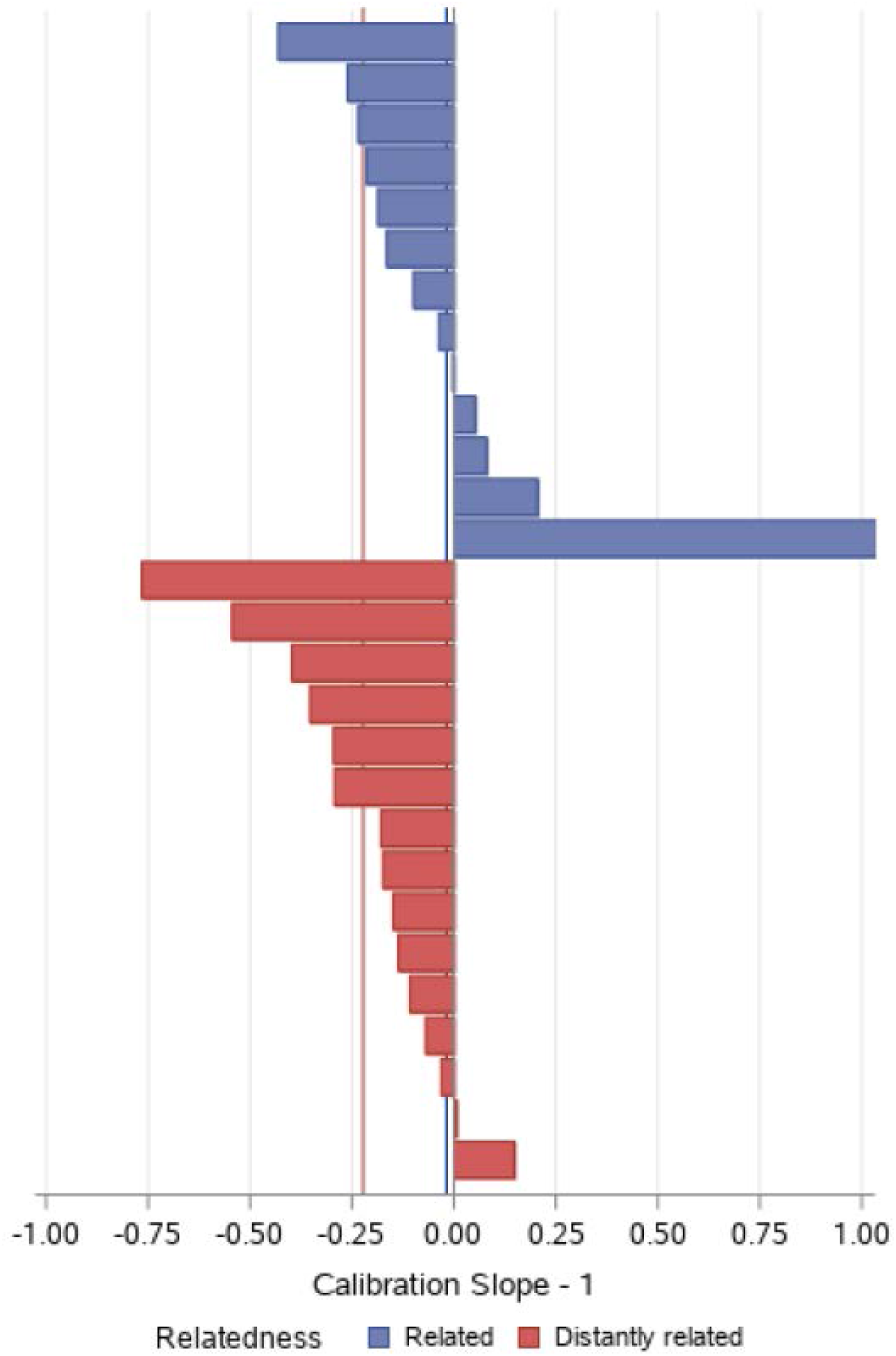

### CPM Calibration in Independent External Validations

The median calibration slope in independent external validations was 0.84 (IQR, 0.72 to 0.98) (Table 2, Figure 3), which indicates some overfitting (i.e. higher than observed predictions in the high risk and lower than observed predictions in the low risk). The E_avg_ standardized to the outcome rate was 0.4 (IQR, 0.3 to 0.8) [i.e. average error was 40% of the observed outcome rate] and standardized E_90_ was 1.0 (IQR, 0.5 to 1.4). For validations done on “related populations” the calibration slope was 0.9 (IQR, 0.79 to 1.05) compared to 0.83 (IQR, 0.65 to 0.93) for “distantly related” matches. For “related matches” the E_avg_ standardized to the outcome rate was 0.3 (IQR, 0.2 to 0.4) and standardized E_90_ was 0.6 (IQR, 0.5 to 1.0) while for “distantly related” validations the E_avg_ standardized to the outcome rate was 0.7 (IQR, 0.4 to 0.8) and standardized E_90_ was 1.2 (IQR 1.0 to 1.7) (Supplement).

### Net Benefit of CPMs

At a threshold set to ½ the observed outcome rate (i.e. prevalence), 32% of CPMs were harmful, 50% were beneficial and 18% would yield outcomes similar to the default strategy of ‘treat none’ (Table 4). With the threshold set to twice the outcome rate, 25% of CPMs were harmful, 54% were beneficial, and 21% would result in outcomes similar to the default ‘treat all’ strategy. At a decision threshold set at the outcome prevalence, 100% of the CPMs were beneficial compared to the default strategies. CPMs tested on ‘distantly related’ populations were more likely to be harmful at thresholds set at ½ the prevalence or twice the prevalence compared to ‘related’ validations (40% vs 23% and 40% vs 8% respectively, Table 5 and 6).

**Table 3.** Effects of Updating

| (n = 28) | Intercept | Intercept + Slope |  | Re-estimation |  |
| --- | --- | --- | --- | --- | --- |
|  | *Compared to Original* | *Compared to Original* | *Incremental Change* | *Compared to Original* | *Incremental Change* |
| **% Change in Discrimination** |  |  |  |  |  |
| c-statistic | N/A | N/A | N/A | 15 (7, 40) | N/A |
| MB c-statistic | N/A | N/A | N/A | 9 (-4, 39) | N/A |
| **% Change in Calibration Error [baseline median standardized E = 0.4 (0.3, 0.8), E90 = 1 (0.6, 1.5)]** |  |  |  |  |  |
| E | -60 (-85, -36) | -76 (-94, -40) | -24 (-67, -7) | -85 (-91, -68) | -29 (-53, 20) |
| E90 | -76 (-83, -43) | -81 (-94, -65) | -23 (-69, -8) | -84 (-92, -76) | -8 (-43, 14) |

**Table 4.** Effects of Updating on Net Benefit (All Matches)

| Validation | Threshold | N | Compared to default strategy |  |  |
| --- | --- | --- | --- | --- | --- |
|  |  |  | % Above | % Neutral* | % Below |
| Original model | Prev./2 | 28 | 50.0 | 17.9 | 32.1 |
|  | Prevalence | 28 | 100.0 | 0.0 | 0.0 |
|  | Prev.*2 | 28 | 53.6 | 21.4 | 25.0 |
| Updated intercept | Prev./2 | 28 | 67.9 | 7.1 | 25.0 |
|  | Prevalence | 28 | 100.0 | 0.0 | 0.0 |
|  | Prev.*2 | 28 | 67.9 | 10.7 | 21.4 |
| Updated intercept and slope | Prev./2 | 28 | 71.4 | 17.9 | 10.7 |
|  | Prevalence | 28 | 100.0 | 0.0 | 0.0 |
|  | Prev.*2 | 28 | 71.4 | 14.3 | 14.3 |
| Re-estimated | Prev./2 | 28 | 89.3 | 3.6 | 7.1 |
|  | Prevalence | 28 | 100.0 | 0.0 | 0.0 |
|  | Prev.*2 | 28 | 100.0 | 0.0 | 0.0 |

**Table 5.** Effects of Updating on Net Benefit (Related Matches)

| Validation | Threshold | N | Compared to default strategy |  |  |
| --- | --- | --- | --- | --- | --- |
|  |  |  | % Above | % Neutral* | % Below |
| Original model | Prev./2 | 13 | 61.5 | 15.4 | 23.1 |
|  | Prevalence | 13 | 100.0 | 0.0 | 0.0 |
|  | Prev.*2 | 13 | 61.5 | 30.8 | 7.7 |
| Updated intercept | Prev./2 | 13 | 84.6 | 0.0 | 15.4 |
|  | Prevalence | 13 | 100.0 | 0.0 | 0.0 |
|  | Prev.*2 | 13 | 69.2 | 23.1 | 7.7 |
| Updated intercept and slope | Prev./2 | 13 | 92.3 | 0.0 | 7.7 |
|  | Prevalence | 13 | 100.0 | 0.0 | 0.0 |
|  | Prev.*2 | 13 | 76.9 | 15.4 | 7.7 |
| Re-estimated | Prev./2 | 13 | 100.0 | 0.0 | 0.0 |
|  | Prevalence | 13 | 100.0 | 0.0 | 0.0 |
|  | Prev.*2 | 13 | 100.0 | 0.0 | 0.0 |

**Table 6:** Effects of Updating on Net Benefit (Distantly Related Matches)

| Validation | Threshold | N | Compared to default strategy |  |  |
| --- | --- | --- | --- | --- | --- |
|  |  |  | % Above | % Neutral* | % Below |
| Original model | Prev./2 | 15 | 40.0 | 20.0 | 40.0 |
|  | Prevalence | 15 | 100.0 | 0.0 | 0.0 |
|  | Prev.*2 | 15 | 46.7 | 13.3 | 40.0 |
| Updated intercept | Prev./2 | 15 | 53.3 | 13.3 | 33.3 |
|  | Prevalence | 15 | 100.0 | 0.0 | 0.0 |
|  | Prev.*2 | 15 | 66.7 | 0.0 | 33.3 |
| Updated intercept and slope | Prev./2 | 15 | 53.3 | 33.3 | 13.3 |
|  | Prevalence | 15 | 100.0 | 0.0 | 0.0 |
|  | Prev.*2 | 15 | 66.7 | 13.3 | 20.0 |
| Re-estimated | Prev./2 | 15 | 80.0 | 6.7 | 13.3 |
|  | Prevalence | 15 | 100.0 | 0.0 | 0.0 |
|  | Prev.*2 | 15 | 100.0 | 0.0 | 0.0 |

### Effects of Updating

The median standardized calibration error (E_avg_) improved from 0.4 to 0.16 [improvement of 60% (IQR 36% to 83%)] with updating of the intercept alone. Calibration error further improved to 0.10 [improvement of 78% (IQR 40% to 94%) over baseline] with updating of the intercept and slope, and 0.06 [improvement of 84% (IQR 63% to 91%)] with re-estimation of the regression coefficients (Table 3). Net benefit generally improved with CPM updating. Sequential updating of the intercept and the intercept and slope appears to offer some protection against harm at the selection thresholds. No CPMs were harmful at two of the chosen threshold values (prevalence and 2*prevalence) following re-estimation of the model regression coefficients (Table 4). When stratified according to relatedness, re-estimation guarded against harm across all decision thresholds for related matches. For distantly related matches, risks of harm at ½ the prevalence persist despite model updating (Table 6).

## Discussion

The primary findings from this study are that ACS CPMs have limited transportability to publically available clinical trial databases. During independent external validations, these CPMs sometimes demonstrated substantial decrements in discriminatory performance, however this was generally due to narrower case-mix (as opposed to model invalidity). More importantly, poor calibration was often substantial, with errors in the predictions that were approximately 40% of the observed outcome rate, such that model use would frequently be expected to cause net harm compared to the best default strategy, as the decision threshold diverged from the prevalence. Model calibration can be substantially improved by updating the model intercept, or the intercept and slope. Using ACS CPMs from the literature to inform decision making when the threshold is far from the outcome prevalence cannot be recommended except in the very unusual circumstance that one knows the CPM is well calibrated to the clinical population under study.

Many potential CPM/ validation database matches were not possible because of missing or alternatively-defined variables in the validation databases. Ultimately, 91% of the ACS CPMs in our Registry could not be matched to the available databases. This finding underscores a major barrier to systematically assessing CPM performance through external validations to comprehensively understand predictive model performance. To overcome this, broader data sharing efforts alone are insufficient; model building efforts should use commonly assessed variables that are collected and defined in a standard fashion in trials and registries.

The decrement in discrimination seen with ACS CPMs mirrors that seen in other disease groups^14^ and there are a number of potential causes of this decrease in performance including overfitting or clinical or regional differences between the derivation and validation populations. The MB-c provides an estimate of model discrimination without any model invalidity and thus enables an assessment of how much of the change in discrimination is due to changes in case-mix. To our knowledge this is one of the first demonstrations using patient-level clinical trial data of the impact case-mix on CPM performance. Generally, there is a more narrow case mix in clinical trial databases compared to registry data or ‘real-world’ populations that are often used to create CPMs.^23^ While it is reassuring that only a small amount of the overall decrement in discrimination is due to model invalidity, the overall observed changes in model performance reinforce the need to understand the similarities and differences between populations when interpreting CPM performance in external validations.

Population relatedness reflects the clinical appropriateness of CPM/validation matches and is associated with CPM validation performance. Expert clinician review of the inclusion and exclusion criteria for each derivation cohort/ validation cohort pairing was required in order to understand to what extent populations represented similar vs. different groups of patients. The appropriateness of potential validations often hinged on subtle clinical differences such has year of enrollment (representing different therapeutic eras) or enrollment timing (emergency room vs inpatient vs post discharge). These clinical details-- with specific attention to characteristics that may make populations dissimilar—should be well understood before attempting to use a CPM since the chance of harm (compared to the default strategy) is higher when populations are distantly related.

We have shown that off-the-shelf CPM risk estimates for patients with ACS can perform poorly, often with significant calibration error, potentially yielding misleading and harmful predictions.^24,25^ Interestingly, relatively simple updating procedures (here merely adjusting for the overall event rate (calibration-in-the-large)) substantially improves the calibration of these predictions. Our previous work has demonstrated that isolated external validations are insufficient to understand broadly how well CPMs perform—since the performance of a given model may vary substantially across different databases—and that the updating that is needed to optimize performance is likely specific to a given cohort.^14^ The required updates for any proposed use case must be understood before CPMs can be used (or trusted) to influence clinical decision-making. Unfortunately, models are used in clinical populations in which calibration to outcome rates may not be feasible.

There are certain limitations to this work. The validation databases generally represent older therapeutic eras since this work reflects databases that are currently available through the BioLINCC program. As a result, the specific validation and updating results presented here are not based on recent trials. While the specific results may not be relevant to inform contemporary clinical decisions for patients with ACS, the approach used here can be applied to modern cohorts to optimize accuracy of predictions. Given the small number of CPM/ validation database matches we were seldom able to match a CPM to more than one validation database (and never more than two). Isolated external validations do not sufficiently describe CPM performance and poor performance here does not necessarily invalidate a given CPM in other settings. We also note that the decision thresholds at which Net Benefit were assessed were arbitrarily chosen; for decision thresholds even further from the prevalence than those examined here, we would anticipate even greater risk of harm. More work is needed to broadly understand performance of these ACS CPMs across various contemporary cohorts.

Our results challenge the concept that using ACS CPMs to inform clinical decisions cannot be worse than the current standard of care. Taking these models from the literature and applying them “off the shelf” to inform decision making risks causing harm. Nevertheless, these findings also point to the potential for improved decision making when models are recalibrated to the populations in which they are applied. This points to the need for a new paradigm for prediction modeling where the best CPMs are identified not by assessing predictive accuracy but instead by assessing clinical utility (i.e. can a CPM actually lead to improved clinical outcomes).^26^ Improving the technological infrastructure to permit continual updating and evaluation of models should be a high priority if we wish to realize the health benefits that better prediction potentially allow.

## Supporting information

Supplemental File- Search Terms

PRISMA checklist

## Data Availability

This study examined data from the Tufts Predictive Analytics and Comparative Effectiveness (PACE) CPM Registry (http://pace.tuftsmedicalcenter.org/cpm).

