## Supplemental File- Search Terms for "The Generalizability of Clinical Prediction Models for Patients with Acute Coronary Syndromes: Results from Independent External Validations"

**Short title:** Generalizability of ACS prediction models

**Keywords:** risk, clinical prediction models, acute coronary syndrome

**Address for Correspondence:**

David M. Kent MD MS

PACE, ICRHPS

Tufts Medical Center

800 Washington Street, Box #63

Boston, MA 02111

**Supplement** Search Terms

((predict$ adj1 model$) or (predict$ adj1 instrument$) or (predict$ adj1 score$) or (predict$ adj1

index)).mp.

((prognos$ adj1 model$) or (prognos$ adj1 instrument$) or (prognos$ adj1 score$) or (prognos$ adj1

index)).mp.

((risk adj1 model$) or (risk adj1 instrument$) or (risk adj1 score$) or (risk adj1 index) or (risk

assessment model or risk assessment instrument or risk assessment score)).mp.

atrial fib$.mp. or exp Atrial Fibrillation/ or exp coronary artery disease/ or exp coronary disease/ or exp

myocardial infarction/ or Myocardial infarct$.mp. or exp Heart Failure, Congestive/ or exp myocardial

ischemia/ or exp cardiovascular diseases/ or exp Cerebrovascular Accident/ or *heart failure/ or *stroke/

or *acute coronary syndrome/

limit 6 to yr="1990 -Current"

**Where current = May 15, 2015 publications

(201205$ or 201206$ or 201207$ or 201208$ or 201209$ or 201210$ or 201211$ or 201212$ or 2013$

or 2014$ or 201501$ or 201502$ or 201503$).ed.
