## Supplemental Table 1 for "The Generalizability of Clinical Prediction Models for Patients with Acute Coronary Syndromes: Results from Independent External Validations"

**Short title:** Generalizability of ACS prediction models

**Keywords:** risk, clinical prediction models, acute coronary syndrome

**Address for Correspondence:**

David M. Kent MD MS

PACE, ICRHPS

Tufts Medical Center

800 Washington Street, Box #63

Boston, MA 02111

**Supplemental Table 1** Heat Map of CPM/Database Matches

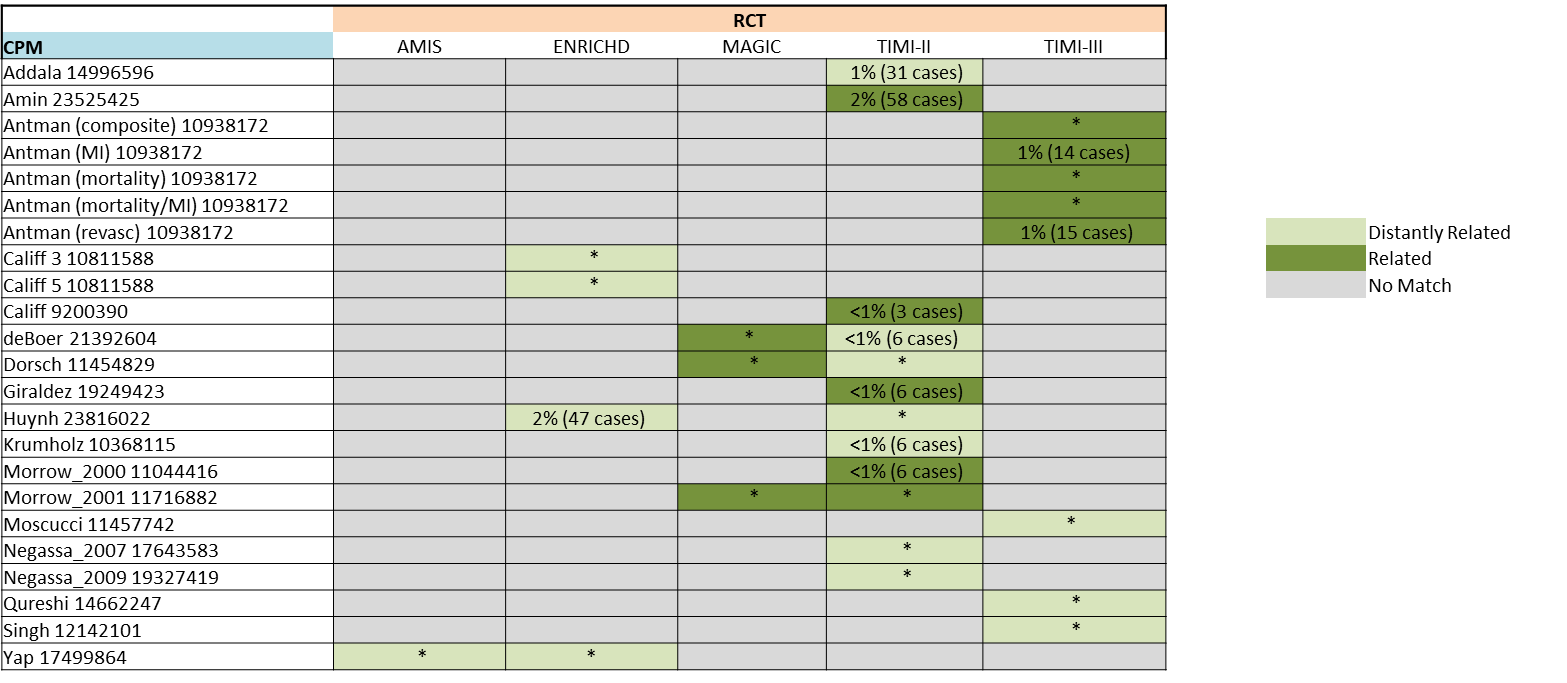

Supplement Table 1. CPM/Database Matches included in this analysis. AMIS is The Aspirin Myocardial Infarction Study, ENRICHED is The Enhanced Recovery in Coronary Heart Disease study. MAGIC is The Magnesium in Coronaries Trial, TIMI-II and TIMI-III are The Thrombolysis in Myocardial Infarction Trials. Light green represents ‘distantly related’ matches. Dark green represents ‘related’ matches. * represents validations done using the full RCT. Otherwise the percentage of vents (number of observations) that are censored because they occurred after the prediction time horizon are shown.
