## Supplemental Table 2 for "The Generalizability of Clinical Prediction Models for Patients with Acute Coronary Syndromes: Results from Independent External Validations"

**Short title:** Generalizability of ACS prediction models

**Keywords:** risk, clinical prediction models, acute coronary syndrome

**Address for Correspondence:**

David M. Kent MD MS

PACE, ICRHPS

Tufts Medical Center

800 Washington Street, Box #63

Boston, MA 02111

**Supplement** Table 2: example of granular relatedness determination.

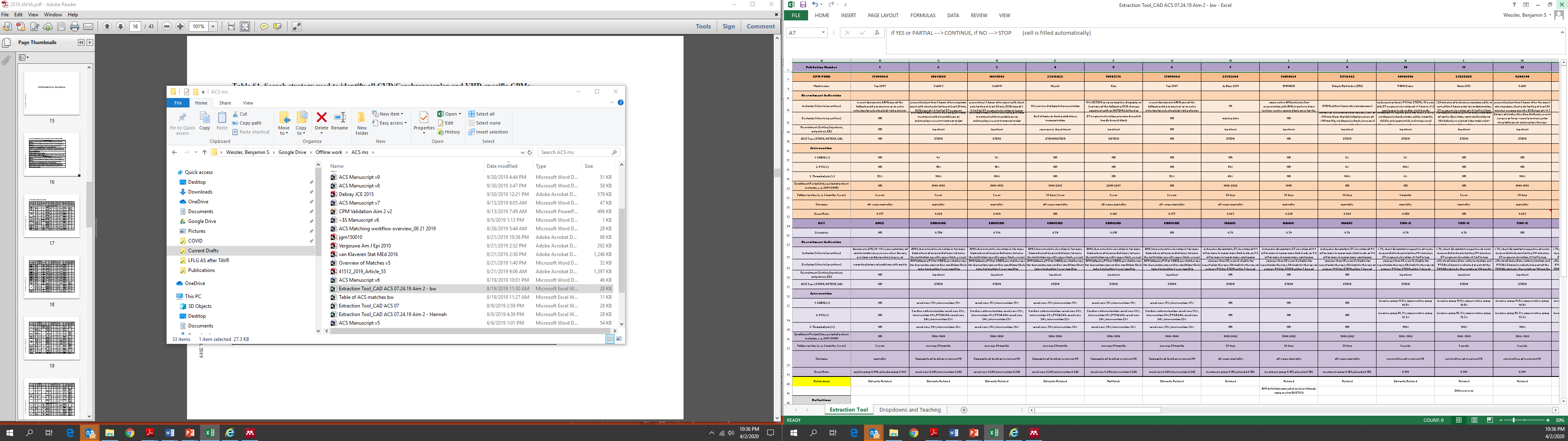
