## Supplemental Table 3 for "The Generalizability of Clinical Prediction Models for Patients with Acute Coronary Syndromes: Results from Independent External Validations"

**Short title:** Generalizability of ACS prediction models

**Keywords:** risk, clinical prediction models, acute coronary syndrome

**Address for Correspondence:**

David M. Kent MD MS

PACE, ICRHPS

Tufts Medical Center

800 Washington Street, Box #63

Boston, MA 02111

**Supplement Table 3** Related Matches

| **(n = 13)** | **Mean (SD)** | **Median (IQR)** | **Range** |
| --- | --- | --- | --- |
| ***Discrimination*** | | | |
| Development c-statistic | 0.72 (0.06) | 0.74 (0.66, 0.76) | 0.63, 0.79 |
| Validation c-statistic | 0.69 (0.04) | 0.7 (0.67, 0.71) | 0.64, 0.77 |
| Validation model-based c-statistic (MBc) | 0.7 (0.06) | 0.7 (0.65, 0.75) | 0.61, 0.8 |
| ***% Change in discrimination due to…*** | | | |
| Total (val.c vs. dev.c) | -14 (25) | -19 (-27, -10) | -41, 49 |
| Case mix heterogeneity (MBc vs. dev.c) | -15 (15) | -14 (-19, -4) | -52, 0 |
| Model validity (val.c vs. MBc) | 3 (33) | -7 (-19, 6) | -34, 76 |
| ***Calibration (8.6% observed outcome rate)*** | | | |
| Slope | 0.98 (0.36) | 0.9 (0.79, 1.05) | 0.57, 2.06 |
| *standardized* E | 0.5 (0.6) | 0.3 (0.2, 0.4) | 0.2, 2.5 |
| *standardized* E90 | 1.3 (2.2) | 0.6 (0.5, 1) | 0.3, 8.4 |
