## Supplemental Table 4 for "The Generalizability of Clinical Prediction Models for Patients with Acute Coronary Syndromes: Results from Independent External Validations"

**Short title:** Generalizability of ACS prediction models

**Keywords:** risk, clinical prediction models, acute coronary syndrome

**Address for Correspondence:**

David M. Kent MD MS

PACE, ICRHPS

Tufts Medical Center

800 Washington Street, Box #63

Boston, MA 02111

**Supplement Table 4** Distantly Related Matches

| **(n = 15)** | **Mean (SD)** | **Median (IQR)** | **Range** |
| --- | --- | --- | --- |
| ***Discrimination*** | | | |
| Development c-statistic | 0.79 (0.04) | 0.78 (0.77, 0.8) | 0.74, 0.9 |
| Validation c-statistic | 0.69 (0.05) | 0.7 (0.66, 0.73) | 0.58, 0.77 |
| Validation model-based c-statistic (MBc) | 0.71 (0.04) | 0.71 (0.69, 0.74) | 0.66, 0.8 |
| ***% Change in discrimination due to…*** | | | |
| Total (val.c vs. dev.c) | -30 (21) | -29 (-44, -15) | -73, 4 |
| Case mix heterogeneity (MBc vs. dev.c) | -23 (13) | -25 (-30, -18) | -46, 7 |
| Model validity (val.c vs. MBc) | -7 (26) | -13 (-28, 14) | -50, 43 |
| ***Calibration (7.9% observed outcome rate)*** | | | |
| Slope | 0.78 (0.23) | 0.83 (0.65, 0.93) | 0.24, 1.15 |
| *standardized* E | 0.6 (0.3) | 0.7 (0.4, 0.8) | 0.1, 1.1 |
| *standardized* E90 | 1.3 (0.7) | 1.2 (1, 1.7) | 0.2, 2.9 |
